# Clonal transitions and phenotypic evolution in Barrett esophagus

**DOI:** 10.1101/2021.03.31.21252894

**Authors:** James A Evans, Emanuela Carlotti, Meng-Lay Lin, Richard J Hackett, Adam Passman, Lorna Dunn, George Elia, Ross J Porter, Mairi H McClean, Frances Hughes, Joanne ChinAleong, Philip Woodland, Sean L Preston, S Michael Griffin, Laurence Lovat, Manuel Rodriguez-Justo, Nicholas A Wright, Marnix Jansen, Stuart AC McDonald

**Author notes:** **Corresponding author:** Dr Stuart McDonald, Centre for Cancer Genomics and Computational Biology, John Vane Science Centre, Barts Cancer Institute, Queen Mary University of London, Charterhouse Square, London, UK EC1M 6BQ. <u>Tel:+44</u>. contributed equally (James A Evans, Emanuela Carlotti). Centre for Inflammation Research, Queens Medical Research Institute, University of Edinburgh, UK. Division of Molecular and Clinical Medicine, School of Medicine, University of Dundee, UK. **Grant support:** This study was funded through a GUTS UK Derek Butler clinical fellowship to JE and a Cancer Research UK programme foundation award to SM (A12446). Institute support was provided through a Cancer Research UK centre grant to the Barts Cancer Centre (C16420/A18066). **Disclosures:** No authors have any conflicts of interest relevant to this manuscript and have all seen and approved this manuscript. **Author contributions:** Study concept and design: SM, NAW, MJ. Acquisition of data and analysis: JE, EC, RJH, MJ, SM. Material support: JC, LD, LL, FH, MG, MM, MRJ. Drafting and critical revision of manuscript: All authors. Funding: SM and JE.

## Abstract

**Background & Aims:** Barrett esophagus (BE) is a risk factor for the development of esophageal adenocarcinoma, however our understanding of how Barrett esophagus evolves is still poorly understood. We demonstrate that dynamic clonal phenotypic changes occur at the gland level, the mechanism by which these changes evolve, and how diversity may play a role in progression.

**Methods:** We analyzed the distribution and diversity of gland phenotype between and within BE biopsies and the background mucosa of those that had progressed to dysplasia or developed BE post-esophagectomy, using immunohistochemistry and H&E analysis. Clonal relationships between distinct gland types were determined by laser capture microdissection sequencing of the mitochondrial genome.

**Results:** Five different non-dysplastic gland phenotypes were identified in a cohort of 64 patients biopsies taken at the same physical location in the esophagus; most non-dysplastic patients showed a single gland phenotype per biopsy, but some showed two or three gland types. We reveal a shared common ancestor between parietal cell-containing oxynto-cardiac glands and goblet cell-containing specialized Barrett glands through a shared somatic mtDNA mutation. We also reveal a similar relationship between specialized and cardiac-type glands, and specialized and Paneth cell-containing glands. The diversity of gland types was significantly increased adjacent to dysplasia compared to non-dysplastic BE and patients with post-esophagectomy BE, suggesting that gland diversity evolves in BE patients over time.

**Conclusions:** We have shown that the range of BE phenotypes represent an evolutionary process and that changes in gland diversity may play a role in progression. Furthermore, we demonstrate common ancestry between gastric and intestinal glands in BE.

**Graphic abstract:** 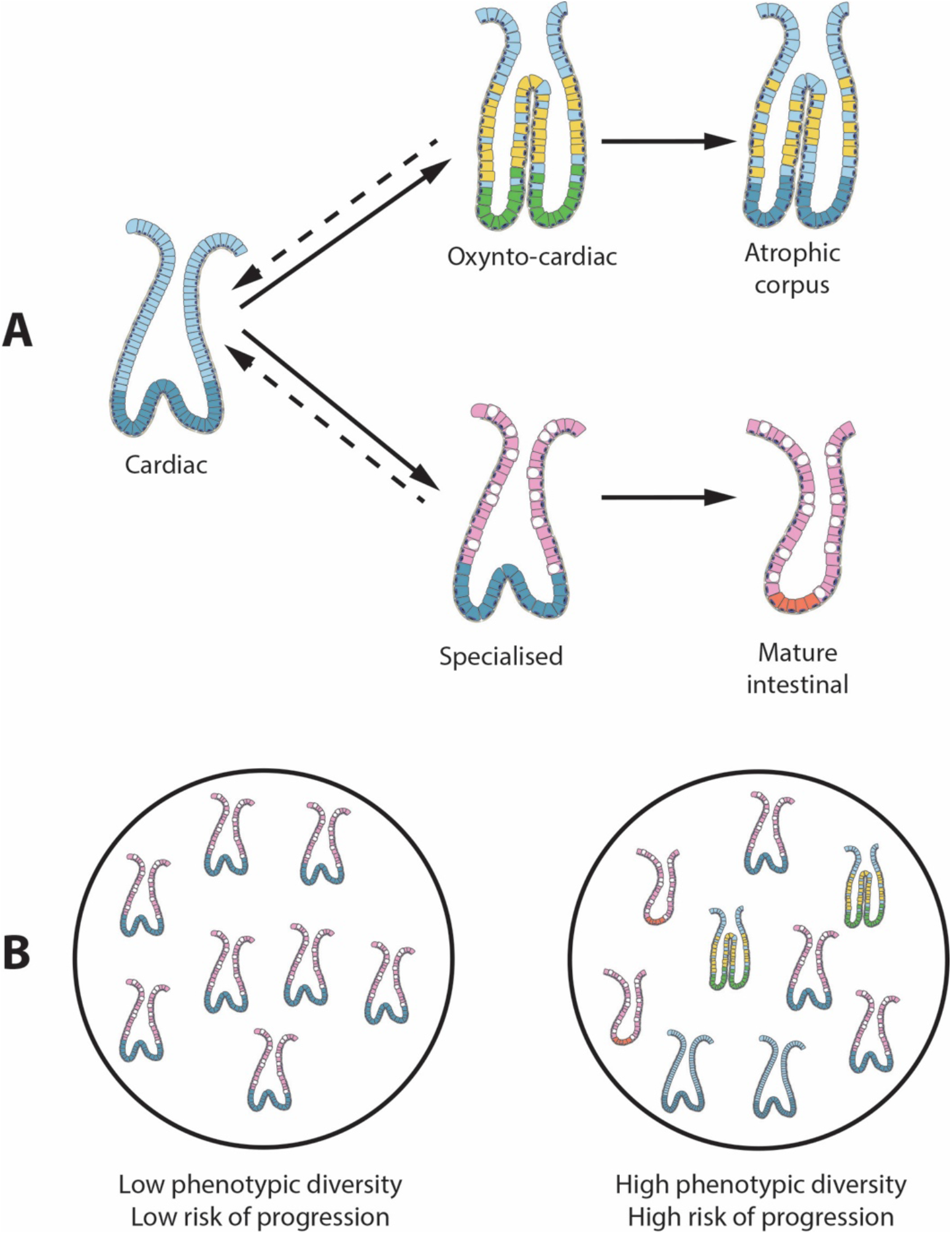

*A) The cardiac gland as the basic unit of Barrett esophagus that can evolve into phenotypes that adapt to the esophageal microenvironment. B) Phenotypic diversity of non-dysplastic glands is associated with the presence of dysplasia or cancer in patients with BE*.

## Background

Barrett esophagus (BE) is the only known precursor condition of esophageal adenocarcinoma (EA) and is characterized by the metaplastic replacement of the normal squamous epithelium of the distal esophagus with a columnar epithelial phenotype that frequently contains intestinal metaplasia (IM)^1^. The diagnosis of BE is based on the histopathologic presence of IM in esophageal biopsies in some countries^2^, but in others only the endoscopic presence of columnar epithelium in the distal esophagus is required^3^. It is widely assumed that the presence of IM assists in stratifying patients’ cancer risk, but there is evidence to suggest this is not always the case^4,5 6^. We have previously shown that glands that do not contain goblet cells can clonally expand, accumulate oncogenic *TP53* mutations and be the source of EA^7^. This finding highlights an important lack of understanding of the evolution of the BE epithelial phenotype^8,9^.

BE displays a rich diversity of morphologically distinct glands^10,11,12^ that contain an admixture of both gastric and intestinal epithelial cell lineages^13^. At present we do not fully understand the scope of epithelial lineage diversity, nor whether these lineages represent an evolutionary pathway which may be altered in patients that progress to dysplasia. Previous studies have documented that genotypic diversity predicts the risk of BE progressing to cancer^14-16^, but it is currently unknown whether this is reflected in phenotypic diversity across the segment. There is controversy as to the mechanism by which genetic diversity evolves with some data showing it is acquired over time^17^ and some demonstrating that diversity is inherent to BE^16^ and is always increased relative to non-progressors^14^. To date, the significance of the evolution of gland phenotype in BE has not been fully appreciated. This is an important omission when we consider that all current diagnoses are entirely based on histopathological analysis^3^ and that natural selection acts fundamentally on phenotype not genotype^18^.

Here we address this unresolved issue by revealing the frequency distribution of gland phenotypes in a cross-sectional BE patient cohort, both at a fixed point within the metaplastic segment (1.0-2.0 cm proximal of the gastroesophageal junction) as well as throughout the BE segment. We then demonstrate ancestral clonal relationships between different gland phenotypes within non-dysplastic BE, indicating phenotypic evolutionary lineages in BE using mitochondrial DNA mutations as clonal marks^19,20^. Finally, we measure gland phenotype diversity in non-dysplastic BE from patients that have never progressed to dysplasia and compare this to non-dysplastic BE adjacent to dysplasia and post-esophagectomy non-dysplastic BE. Together this data demonstrates, for the first time, the phenotypic evolutionary pathway within BE.

## Methods

### Patients

Patients were recruited from the surveillance BE endoscopic clinic at Barts Health NHS Trust and from the archives of both the Royal London Hospital and University College London Hospital approved under multicenter ethical approval from London research ethics committee (11/LO/1613 and 15/LO/2127). Post-esophagectomy BE esophagus specimens were accessed under County Durham and Tees Valley 2 research ethics committee approval (08/H0908/25) and from University College London hospital (as per above). Snap frozen biopsies and formalin-fixed paraffin-embedded (FFPE) specimens were used in this study.

#### Cohort 1a

A series of 64 biopsies from 51 BE patients were collected from 1.0-2.0cm proximal of the gastroesophageal junction and were FFPE-preserved. All biopsies met the following inclusion criteria: 1) Biopsies taken at the same anatomical height within the esophagus, regardless of BE maximum length; 2) Taken from the BE lesion identified during endoscopy, and 3) Absence of dysplasia or cancer at the time of endoscopy or any previous history of dysplasia. The mean age of the patients within cohort 1 was 62.2 (range 27-89) years, the female to male ratio was 1:4.9 and the mean maximum BE segment length was 4.5 cm (range 1.5-14 cm, median = 4.0 cm). For 25 of these patients, we obtained further archival FFPE H&E sections from all biopsies taken at the same surveillance endoscopy.

#### Cohort 1b

Fresh frozen biopsies taken from 10 patients in cohort 1a (adjacent biopsies) during the same endoscopy.

#### Cohort 2

FFPE-preserved 99 BE biopsies (19 patients) showing no dysplasia or history of dysplasia and (ii) 21 endoscopic mucosal resection (EMRs) specimens (18 patients) for high grade dysplasia with adjacent non-dysplastic BE (age range 43-65 years).

#### Cohort 3

31 biopsies from 19 patients with post-esophagectomy BE (neo-BE). All patient biopsies were taken a minimum of 2 years post-esophagectomy for adenocarcinoma. No data was collected for patient age or BE length. All samples were FFPE.

### Gland phenotype identification

Two independent experienced pathologists determined gland phenotype in FFPE and frozen BE tissue sections (MJ & NAW) by identifying the presence of either chief cells, parietal cells, goblet cells, foveolar cells or Paneth cells by H&E and by immunohistochemistry (IHC) analysis (**Supplemental figure 1**).

### Immunohistochemistry

Serial 5μm FFPE tissue sections were dewaxed in xylene and hydrated through a graded ethanol series to water. Antigen retrieval was performed in boiling Tris-EDTA pH 8.0 (Sigma, Poole, UK) or sodium citrate pH 6.0 (FisherChemicals, Loughborough, UK) for 10 minutes depending on each primary antibody (**Supplemental Table 1**). Endogenous peroxidase activity was blocked with 3% hydrogen peroxide (FisherChemical, UK) for 10 minutes followed by Protein Block, Serum-Free solution (Agilent Technologies Ltd, Stockport, UK) for 30 minutes. No endogenous alkaline phosphatase was detected.

Double IHC was performed in a specific sequence on serial sections of H^+^/K^+^-ATPase (Horseradish peroxidase-diaminobenzine (HRP-DAB)) and Pepsinogen (alkaline phosphatase (AP)-Blue) (set 1) and for MUC5AC (HRP-DAB) and MUC2 (AP-Blue) (set 2) and Defensin 6α only (AP-Blue)(set 3) (**Supplemental figure 1**). Staining was performed using a secondary-biotinylated antibody followed by a tertiary streptavidin conjugated with HRP. For HRP antibodies, each was diluted in Ready-to-Use diluent (Agilent, UK) and incubated for 1 hour at room temperature (RT) followed by incubation with either a goat anti-mouse IgG (Agilent, UK) or swine anti-rabbit (Sigma) at a 1:200 dilution for 45 minutes at RT depending on the primary antibody (**Supplemental table 1**). Streptavidin HRP (Agilent, UK) was then added (1:100) and incubated for 30 minutes and DAB Peroxidase substrate was added until a brown color developed (Vectro Labs Ltd, Peterborough, UK). This was followed by primary antibodies for AP-Blue, a secondary-biotinylated antibody, then tertiary streptavidin conjugated to AP. Vector blue substrate (Vector Labs Ltd, UK) was then added using the same protocol described for HRP. The same protocol was followed for frozen sections with the exception of not performing antigen retrieval and section thickness was 10•m.

### Laser-capture microdissection

Serial 10μm frozen sections were cut onto P.A.L.M. membrane slides (Zeiss, Germany) previously treated with UV exposure for 30-40 mins. To delineate gland outline in frozen material, sections were subjected to dual enzyme histochemistry for cytochrome *c* oxidase and succinate dehydrogenase as per previously published protocols^19,21^. In all cases, sections were left to dry, then microdissection was performed using a P.A.L.M laser dissection microscope (Zeiss, Germany). Microdissected cells were digested in 14μl Picopure digestion buffer (Life Technologies, UK) at 65°C for 3 hours, followed by proteinase K inactivation at 95°C for 5 mins.

### Mitochondrial PCR sequencing

A nested PCR protocol was used as previously published^19^. Briefly, the mitochondrial genome from each microdissected area was amplified into nine, 2 kb fragments, which were subsequently re-amplified into 500 bp fragments. Primer sequences and PCR conditions were used as previously described^19^. The second round PCR primers contained an M13 sequence to facilitate sanger sequencing. PCR products were ExoSaP-treated according to manufacturer’s protocol (GE Healthcare, UK) and Sanger sequenced by Eurofins Genomics (Ebersberg, Germany). Obtained sequences were viewed using 4Peaks software (https://nucleobite.com) and compared to the revised Cambridge reference sequence using online tools provided at www.mitomap.com. Polymorphisms and non-epithelial mutations were eliminated from analysis by comparison with sequences from a microdissected area of stroma. Each mutation was confirmed using the same PCR sequencing protocol repeated from the original DNA sample.

### Statistics

Statistical analysis was performed using a one-way ANOVA (Kruskal-Wallis) test for assessing phenotype distribution. An unpaired Student’s t-test or a Mann-Whitney U test was used when comparing BE length and diversity, changes in diversity and changes in phenotypes where data was either normally distributed or not, respectively.

Diversity was measured using both a richness score (a record of the number of gland types visible) and the Shannon index (SI)^15^ that takes into account the number of gland types and their relative frequency within a tissue specimen. The SI was calculated as;

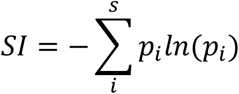

Where *s* is the number of species, pi is the frequency of each gland phenotype (*i*) within a tissue specimen; SI was calculated as the sum of the natural log (*ln*) of every *p*_*i*_ (*ln(p*_*i*_)). Significance was determined using a two-sample Mann Whitney U test.

## Results

### Identification and distribution of different gland phenotypes in BE

Here we provide a detailed analysis of gland phenotype from cohort 1 of BE patients (described in the materials and methods). Figure **1A** shows representative H&E FFPE sections of 5 histologically confirmed phenotypes detected in our BE cohort. We additionally developed a lineage-specific expression profile using IHC to assist in identifying gland phenotype. Gland species were identified as: atrophic corpus (H^+^K^+-^ATPase^+^/Pepsinogen^+^), oxynto-cardiac (H^+^K^+^ATPase^+^/Pepsinogen^-^), simple cardiac type (MUC5AC^+^, MUC2^-^) specialized BE (MUC5AC^+^, MUC2^+^), and mature intestinal (MUC5AC^+/-^, MUC2^+^, Defensin6α^+^ or HD6^+^) (**Supplemental figure 1**).

**Figure 1.**
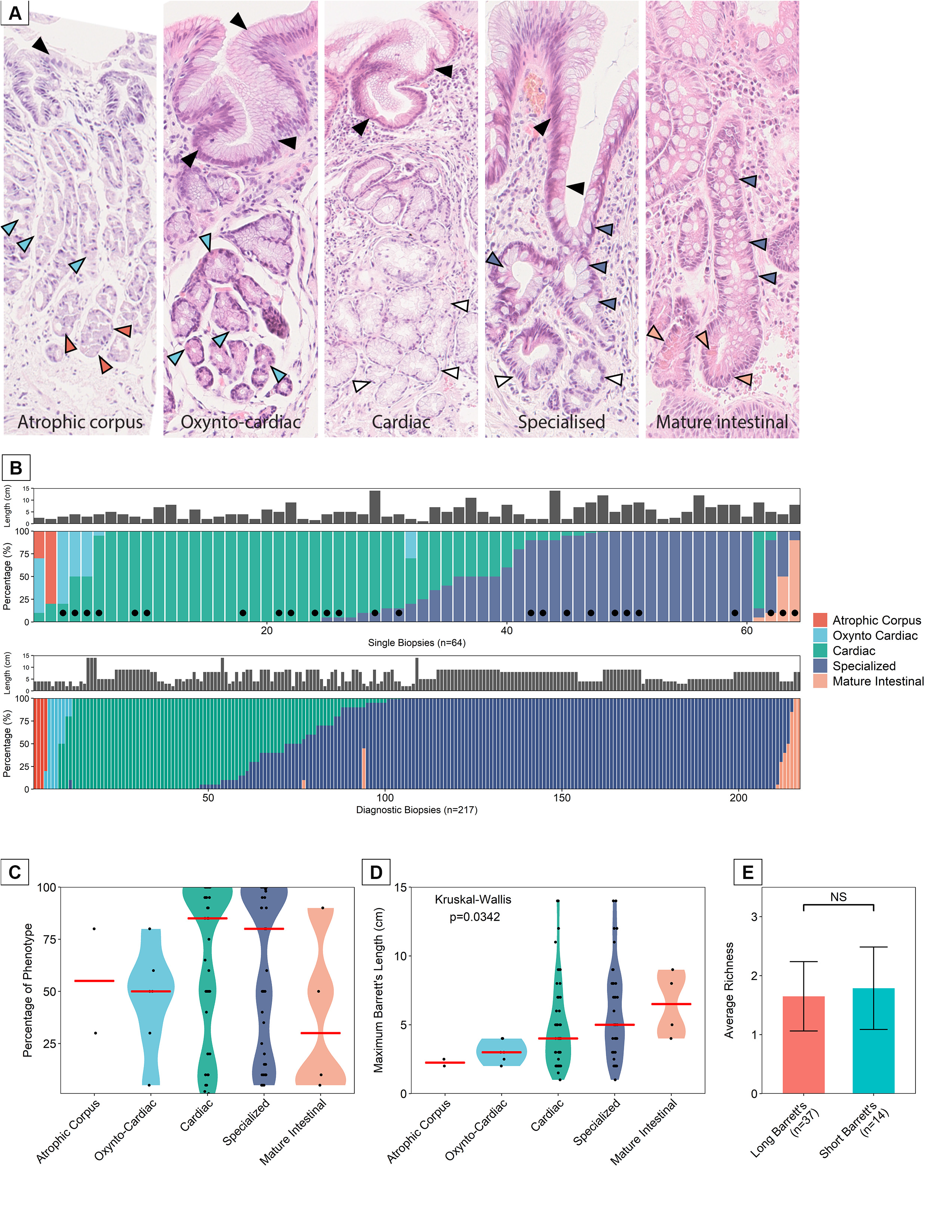
The histological gland phenotypes observed in BE. (**A**), from left to right: Atrophic corpus glands containing parietal cells (Δ), chief cells(Δ) and foveolar cells(Δ); oxynto-cardiac glands containing parietal cells(Δ), foveolar cells (Δ) but not chief cells; cardiac-type glands containing foveolar (Δ) and mucous secreting cells (Δ) only; specialized glands containing goblet cells (Δ), foveolar cells(Δ) and mucous secreting cells (Δ); mature intestinal glands containing goblet cells (Δ) and Paneth cells (Δ). (**B, top**) Distribution of gland types within single biopsies taken from 1.0-2.0 cm proximal of the gastroesophageal junction in observable salmon-pink mucosa. (**B, bottom**) Distribution of gland types throughout the BE lesion from biopsies taken throughout those in (B, top) marked (•). (**C**) A summary of the frequency of gland types through all the single biopsy cohort. (**D**) Relationship between specific gland types and the maximum length of the BE lesion. (**E**) There is no difference in the number of unique gland types per biopsy between long and short segment BE.

The distribution of each gland phenotype in a cohort of 64 biopsies (from 51 patients, 10 of whom had follow-up biopsies) where a single biopsy was taken at 1.0-2.0 cm above the gastroesophageal junction is shown in **Figure 1B (top)** and the corresponding diagnostic biopsies taken during the same endoscopy where available (marked as •) are shown in **Figure 1B (bottom)** (n=25 patients, 217 biopsies). The same overall phenotype pattern is observed in the single and the overall diagnostic biopsies. Individual patient phenotype data based on location of diagnostic biopsies within the BE lesion are presented in **Supplemental figure 2**. From this it is clear that the previously reported distribution of gland phenotype from proximal to distal ends of the lesion^10^ is not apparent in the majority of cases, although this may be due to differences in cohort or overall Barrett segment length. There is a clear dominance of cardiac and specialized gland types from the single biopsy cohort (**Figure 1B and C**) as well as from the diagnostic biopsy cohort (**Figure 1C**). There was a positive relationship between the overall size of the lesion and the gland types detected at 1.0-2.0cm, where the longer BE segment was more likely to show specialized and mature intestinal glands (**Figure 1D**). This finding is in line with a previous report^22^. Most single biopsies contained either one or two gland types (n=30 (46.9%) and n=30 (46.9%) respectively). In a small number of cases, 3 phenotypes were observed within single biopsies (n=3 (4.7%)) and these displayed atrophic corpus or mature intestinal glands. Interestingly, the presence and diversity of specific gland phenotypes from single biopsies at 1.0-2.0cm was not dependent on BE lesion maximum length (**Figure 1D**), nor was it dependent on patient age (**Supplemental figure 3**). Phenotypic richness was not associated with patient age, segment length, or the number of biopsies taken at any single endoscopy as per bootstrapping analysis (**Supplemental figure 4**). Additionally, 10 patients with follow-up over time data available (n=5 (50%)), show no change in diversity, although this is highly variable (**Supplemental figure 5**).

### Evolution of gland phenotypes in BE

These analyses underscore significant phenotypic heterogeneity, even in biopsies from uncomplicated Barrett’s segments. Multiple phenotypes in a single biopsy may be a consequence of independent parallel evolution, or, alternatively, shared branching evolution. To investigate phenotypic gland evolution in BE, we determined if divergent gland phenotypes within biopsies share a common ancestor. In total, 10 snap frozen biopsies from cohort 1b that demonstrated at least two gland types, as verified by IHC, were subjected to LCM mtDNA sequencing. Glands of each individual phenotype were microdissected. Of these cases, 6 did not show any mtDNA mutations with sufficient heteroplasmy to be used as a clonal marker. Of the remaining four cases, one case (H&E **Figure 2A)** showed the presence of both H^+^/K^+^-ATPase parietal cell-containing oxynto-cardiac glands (**Figures 2B** and **2Di** at high power) and MUC5AC^+^/MUC2^+^ goblet cell-containing specialized glands (**Figures 2C** and **2Ei** at high power). Pre- and post-LCM micrographs are shown (**Figures 2Dii,iii** and **2Eii,iii**). A common somatic *m*.*303-311 Cins* mtDNA mutation in the H-strand replication origin region (*MT-OHR*)(**Figure 2E & F**) was observed between the entire parietal cell-rich area and 4 out 5 MUC2^+^ glands. This mutation was not present in the stroma and was also not present in one additional MUC2^+^ gland within the same biopsy, indicating the presence of multiple clonal lineages within phenotypes. Furthermore, morphological examination of an H&E from a cardiac biopsy from this patient did not show presence of goblet cells. This shows for the first time, a clonal relationship between gastric body-type glands and specialized glands in BE.

**Figure 2.**
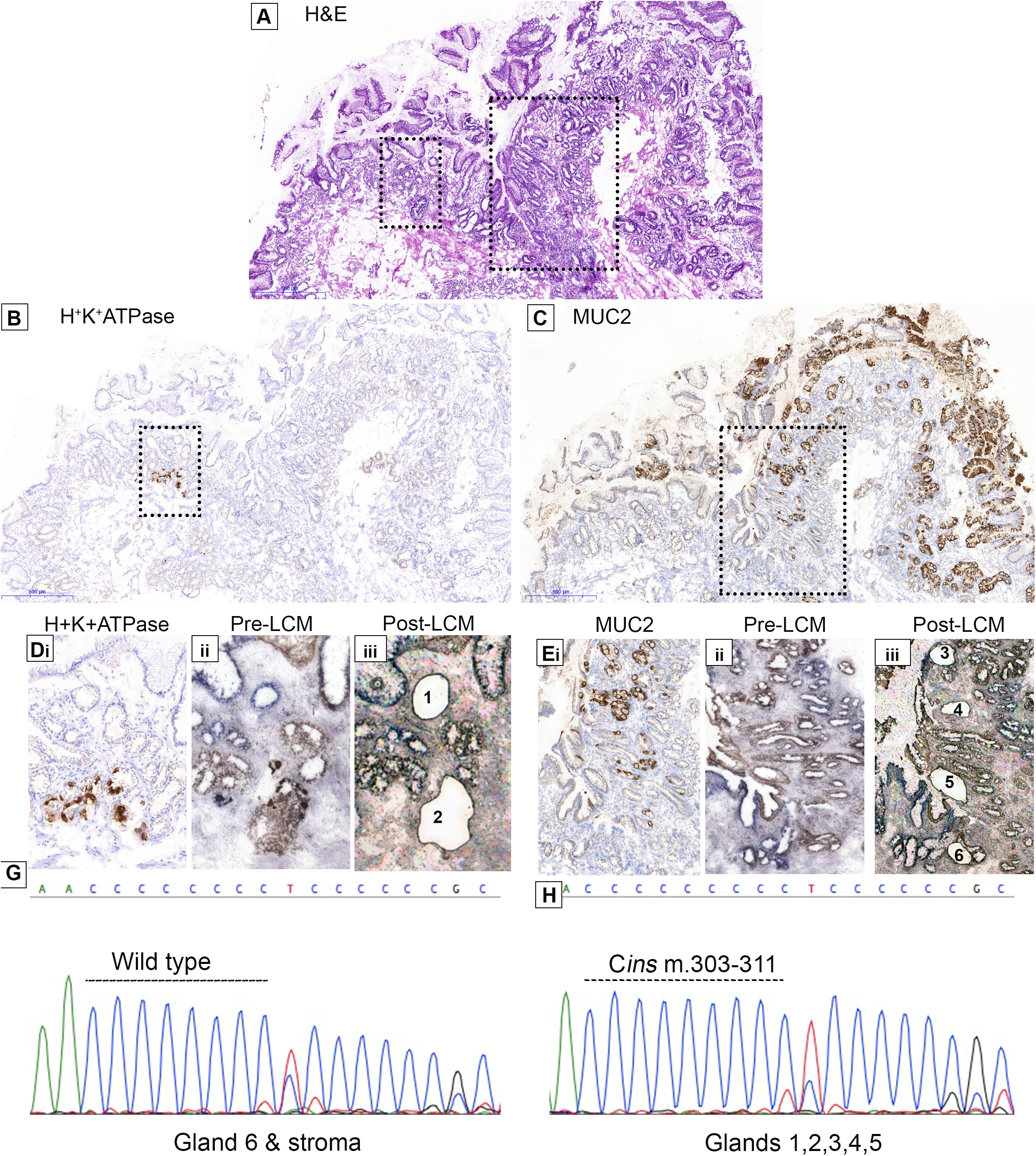
A common ancestry between oxynto-cardiac and specialized epithelium in BE. (**A**) H&E of a BE biopsy showing parietal cells and goblet cells. (**B**) H^+^K^+^ATPase^+^ glands and (**C**) MUC2^+^ glands on the same section. (**Di-iii**) and (**Ei-iii**) shows high power magnification images of (**C**) and (**D**) respectively with pre- and post-microdissection images. PCR sequencing revealed a common, complex *m*.*303-311 Cins* mutation in the *MT-OHR* region of the mt genome present in both MUC2^+^ and H^+^K^+^ATPase^+^ glands but not in stroma and in one other gland sequenced (**G &H**).

The most common gland phenotypes observed in cohort 1 were cardiac and specialized BE glands. To determine if these can show a common ancestor, we investigated a frozen biopsy (**H&E, Figure 3A**) wherein all glands bar a single gland were MUC2^+^ (**Figure 3B**). MUC5AC (**Figure 3C**) staining revealed a mixture of positive and negative glands, indicating the presence of specialized and mature intestinal glands. LCM of the single MUC2^-^ MUC5AC^+^ cardiac-type gland and a neighboring MUC2^+^ MUC5AC^-^ mature intestinal gland (**Figure 3D&E**) revealed a common *m*.*10492 T>C* mutation in the *MT-ND4L* region (**Figure 3F**) that was not present elsewhere in the biopsy (**Figure 3H**).

**Figure 3.**
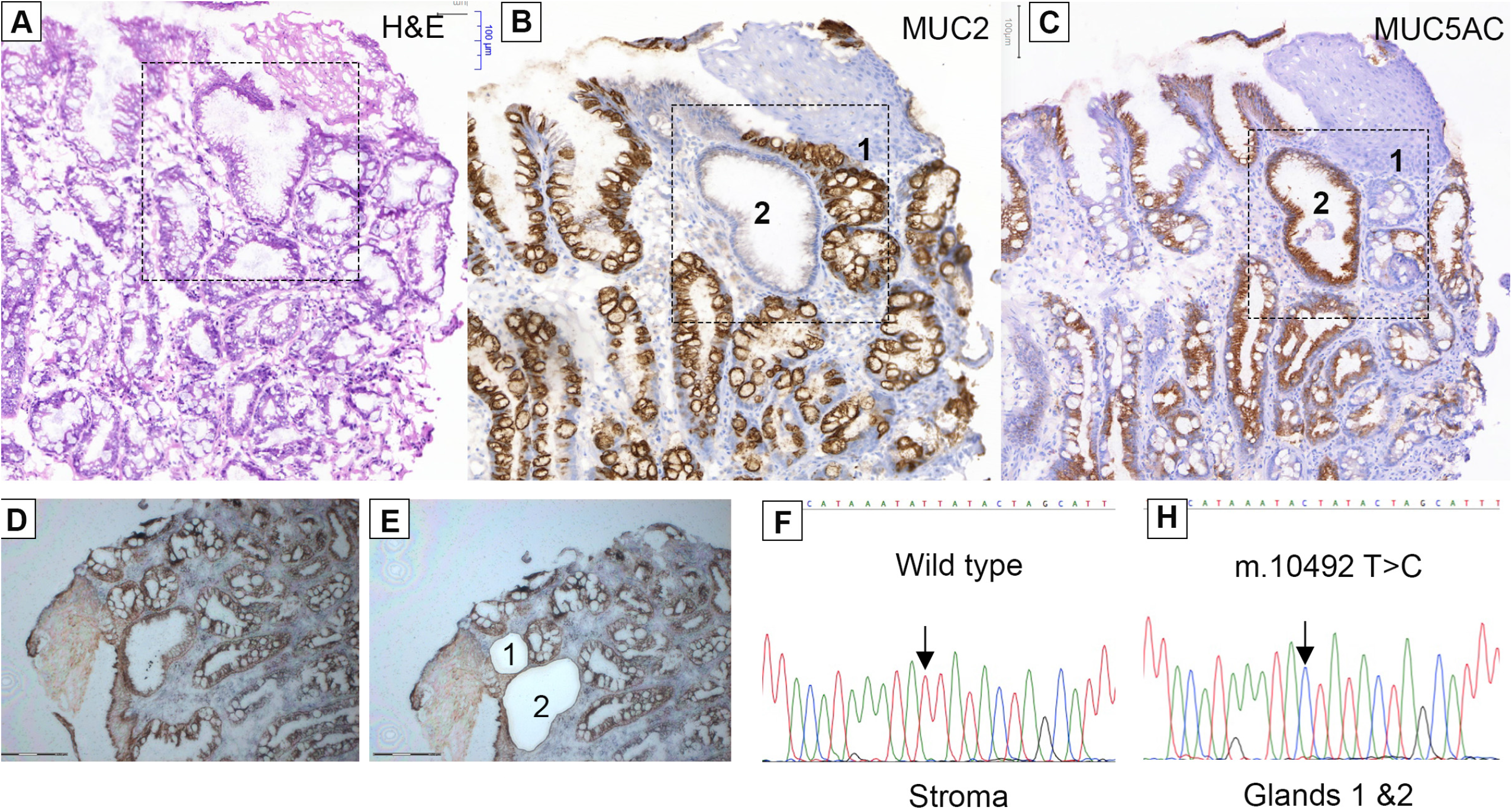
A common ancestry between cardiac-type and mature intestinal epithelium. (**A**) H&E of a BE biopsy containing both cardiac and mature intestinal epithelium (highlighted region). (**B**) MUC2^+^ and MUC2^-^ and (**C**) MUC5AC^+^ and MUC5AC^-^ glands present. (**D&E**) Pre- and post LCM images respectively. Gland 1 is a MUC2^-^ MUC5AC^+^ cardiac type gland. Gland 2 is a MUC2^+^ MUC5AC^-^ gland. (**F&H**) Glands 1&2 revealed a common *m*.*10492T>C* mutation in the *MT-ND4L* region of the mt genome not present in stroma and other glands.

In a separate patient we observed the presence of specialized (MUC2^+^MUC5AC^+^HD6^-^) and mature intestinal glands (MUC2^+^ HD6^+^) (**Figures 4A** H&E, **4B** HD6, **4C** MUC5AC and **4D** MUC2 respectively) and **Figures 4Ei-x** at higher power). We microdissected 7 glands in total (3 HD6^+^, 4 HD6^-^). We detected two somatic mtDNA mutations (*m*.*2283 T>C* and *m*.*2217 T>C* both located in the MT-RNR2 region) in 2 HD6^+^ and 2 HD6^-^ glands suggesting a common ancestor between the two gland lineages (**Figures 4Fi & ii**), not present in one other HD6^+^ and one other HD6^-^ gland (**Figures 4Fiii&iv**). This demonstrates clonal phenotypic evolution from specialized to mature intestinal type glands which is strongly suggestive that this is occurring independently on more than one occasion within the same biopsy. **Supplemental Figure 6A-D** demonstrates another case showing the same relationship between cardiac, specialized and mature intestinal glands revealing a common *m*.*7148 C>T* mutation in the *MT-C01* region of the mt genome. Together these data show that gland phenotypes are not fixed and that dynamic changes in gland phenotype are clonal transitions.

### Phenotypic diversity in non-dysplastic BE adjacent to dysplasia

Overall, these data show the diversity, clonal relationship and ordering of phenotype evolution in non-dysplastic BE. Our analysis suggests that phenotypic transitions are bottleneck events. Successive bottlenecks increase the risk of progression. Therefore, we hypothesized that BE adjacent to dysplasia would demonstrate greater evidence of diversity. To determine if gland phenotype is altered in BE progression to dysplasia or cancer we investigated an additional group of patients (Cohort 2; see *materials and methods*) of FFPE archived material of 99 biopsies, from 19 patients with non-dysplastic BE and no previous history of dysplasia or cancer, 33 EMRs from 18 patients with confirmed dysplasia/cancer that also displayed surrounding non-dysplastic BE, and 19 patients (31 biopsies) that developed post-esophagectomy BE (Cohort 3, neo-BE) within 2 years of complete removal of both the cancer and any remaining BE. **Figure 5A** shows a representative H&E section of an endoscopic resection specimen revealing non-dysplastic BE (white dashed square) adjacent to BE neoplasia (intramucosal adenocarcinoma, green dotted circle). It is clear from this section that there are multiple gland phenotypes in the non-dysplastic BE adjacent to BE neoplasia and at high power magnification, it is clear that these are distinct glands (**Figure 5A insert**). Representative H&E images from neo-BE patients are shown in **Figure 5B&C**).

**Figure 4.**
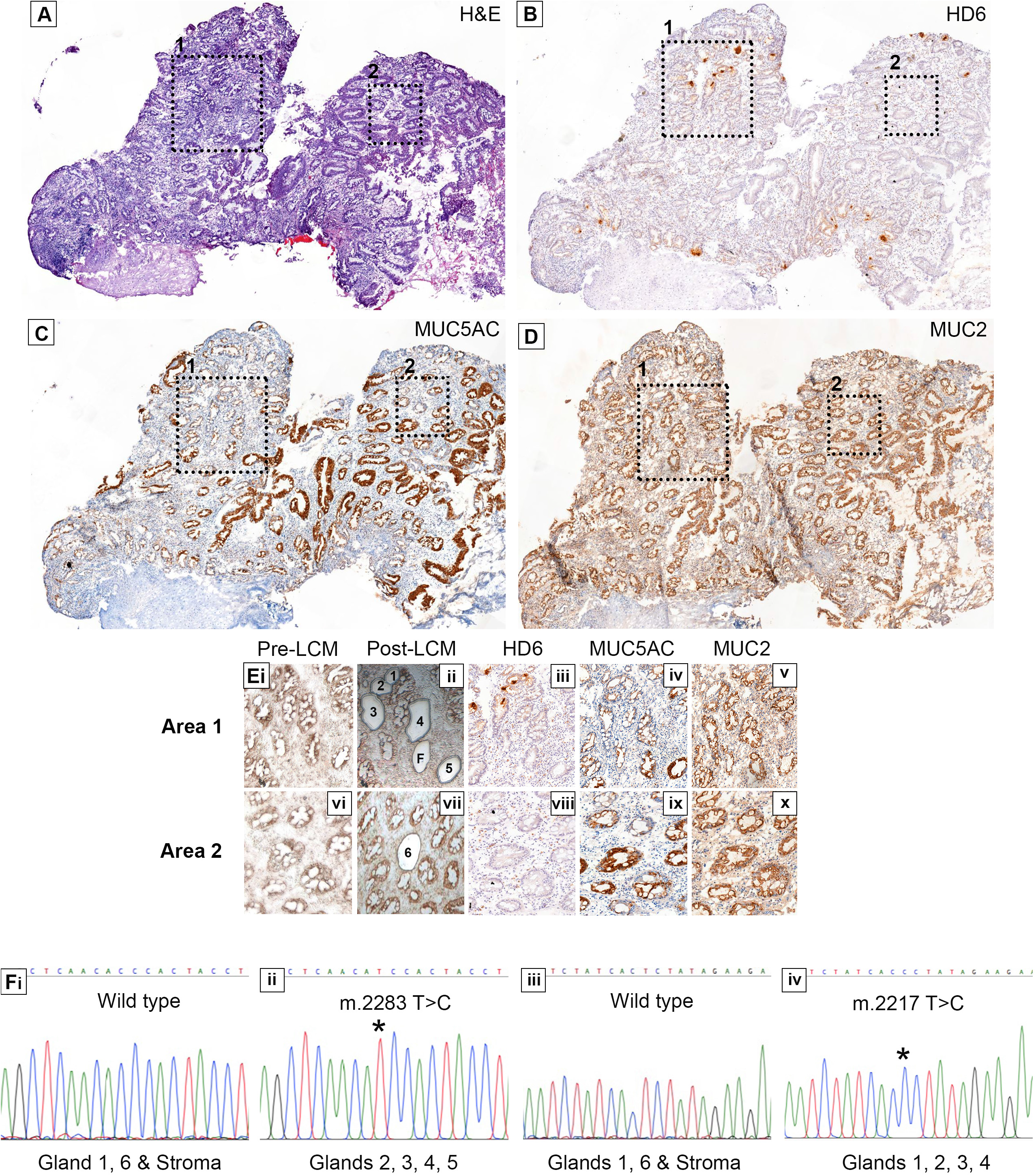
The evolution of specialized and mature intestinal glands in BE. (**A**) H&E of a biopsy containing both gland phenotypes confirmed by IHC for (**B**) Defensin6α^+^ (HD6^+^) and HD6^-^ glands, (**C**) MUC5AC^+^ and MUC5AC^-^ glands and (**D**) MUC2+ glands. Dash-lined boxes indicate areas of interest. (**Ei-x**) Shows the two areas of interest at high power with pre- and post-LCM images as well as those of mucin and defensin staining of each area: In total, 7 glands were microdissected (3 HD6^+^, 4 HD6^-^, one of which failed amplification marked ‘F’). We observed two somatic mt DNA mutations in 4 out 6 glands (*m*.*2283 T>C* and *m*.*2217T>C*) in the *MT-CO1* region of the mt genome containing both specialized and mature intestinal glands that were not present in 2 HD6^+^ glands and the surrounding stroma (**Gi-iv**).

**Figure 5.**
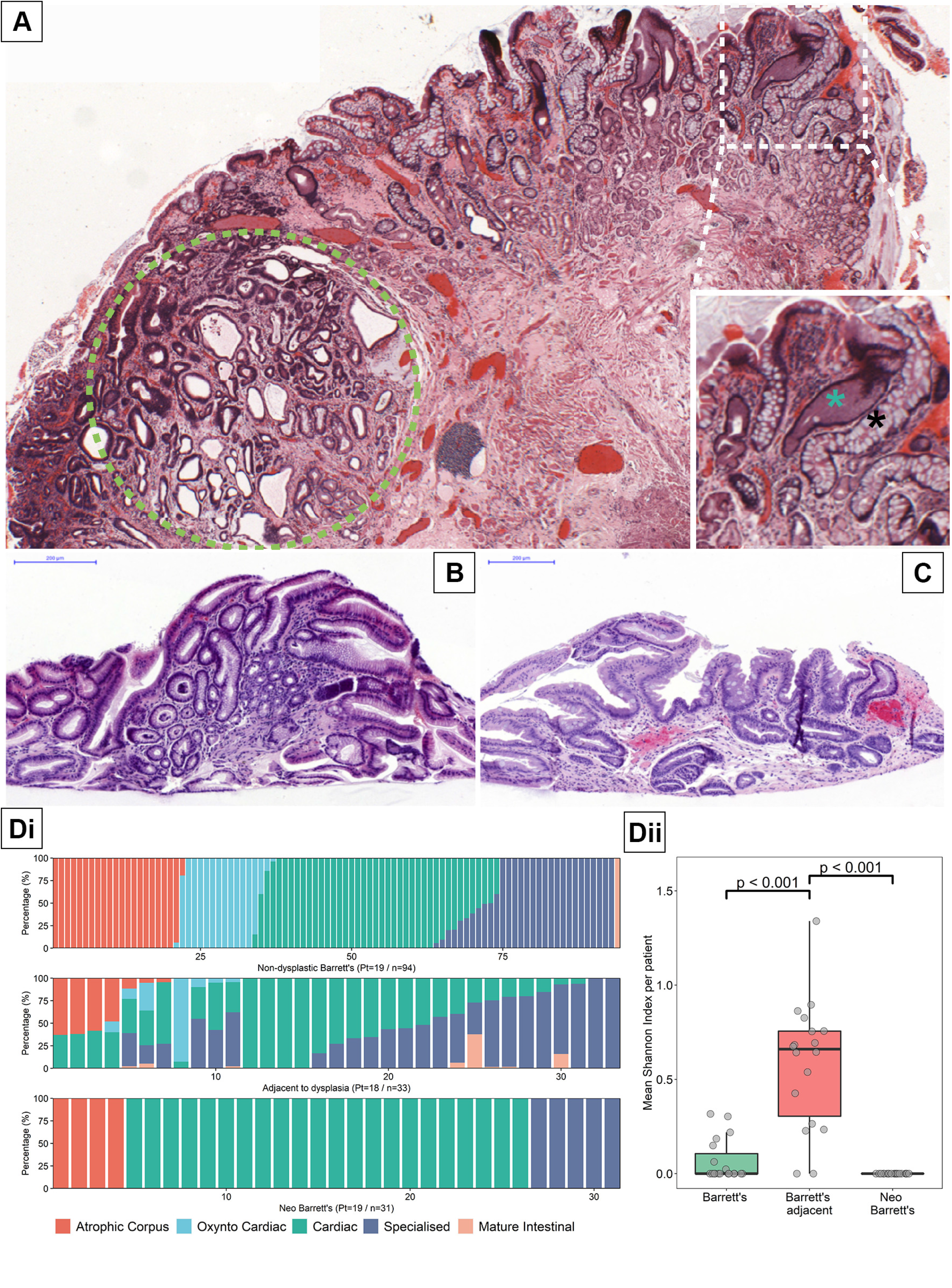
BE gland phenotype diversity is associated with dysplasia. (**A**) An endoscopic mucosal resection H&E showing an area of high grade dysplastic (green dotted line) with surrounding non-dysplastic BE showing the presence of multiple non-dysplastic gland phenotypes. Insert shows a distinct cardiac type gland adjacent^8^ to each other. (**B**) and (**C**) post-esophagectomy BE (neo-BE) biopsies showing cardiac-type epithelium and specialized epithelium. (**Di**) The Shannon index of 99 biopsies (from 25 patients with never-to-dysplasia BE, 32 pieces of EMRs from 18 patients showing BE adjacent to dysplasia and 20 neo-BE cases showing that non- dysplastic BE adjacent to dysplasia is significantly more diverse than either never-to-dysplasia or neo-BE (P<0.001). (**Dii**) The mean Shannon Index per patient (P<0.001). (**Eiii**) The mean Shannon Index of per patient data normalized to 100 glands (P<0.001). In each analysis no diversity was observed in the neo-BE cases.

We then compared the phenotypic diversity of BE, BE adjacent to dysplasia and neo-BE using a biopsy per biopsy approach (**Figure 5Di**) The Shannon Index (see materials and methods) based on the percentage of epithelium of a given gland phenotype within each specimen, was used as a means to compare phenotypic diversity between the 3 groups (**Figure 5Dii**). Interestingly, we find that BE adjacent to dysplasia is significantly more phenotypically diverse compared to BE biopsies from patients without dysplasia or neo-BE (**Figure 5Dii**). Individual biopsies capture a smaller mucosal surface area compared to shoulder regions of EMR specimens which may underestimate diversity in the biopsy group. To address this, bootstrapping analysis was performed on all of the biopsies in our cohort (**Supplemental materials and methods**) which showed that randomly sampling a larger number of glands from these biopsies would not alter the outcome of Shannon diversity (**Supplemental Figure 7**). Neo-BE biopsies showed no per-biopsy diversity whatsoever and the dominant phenotype observed was cardiac (n=22/31 biopsies in 13/19 patients) as has been observed before^23^. Specialized and corpus type glands were identified in a minority of patients (n=5/31 biopsies in 4/19 patients and n=4 biopsies in, n=4 patients respectively). Two Neo-BE patients showed more than one phenotype (**Supplemental Figure 8**) but in distinct biopsies only. A patient-per-patient biopsy phenotype distribution for all three groups is shown in **Supplemental Figure 9**. This data shows that areas of non-dysplastic BE adjacent to dysplasia is significantly more diverse and perhaps could act as a potential predictive biomarker for progression in surveillance biopsies. Therefore, the lack of diversity in neo-BE cases, which can be considered a model of early BE, may suggest that phenotypic diversity is acquired over time.

## Discussion

The study of gland phenotype in BE has been largely overlooked^10-12^. Considering that all BE diagnoses are based on phenotypic observations, there is an unmet need to determine the range, evolution, mechanism and diversity of these phenotypes. Our cohorts appear to reflect those included in previous studies by identifying similar gland types^10^, but with a much broader interpretation of phenotype and their diversity. The ancestral relationships we uncover between gland phenotypes provides important information about the evolution of BE to cancer. We show that evolution of BE can involve a change of phenotype at the stem cell level (niche succession^24,25^). Importantly, we show that a mature gastric gland phenotype expressing parietal cells shares a common ancestor with specialized BE indicating a phenotypic adaptation of BE along a gastric differentiation presumably as a result of natural selection acting on gastric phenotypes. We show that multiple phenotypic changes represent clonal phenotypic evolution within BE and that an increase in their diversity is associated with progression. However, *de novo* post-esophagectomy BE does not show any within biopsy diversity and this suggests that gland phenotype diversity is acquired over time and may not be present during the early development of BE.

We observe five well-defined gland phenotypes in our cohort of biopsies taken at the same anatomical location within the esophagus (1.0-2.0 cm proximal of the gastric folds). Of note, we did not observe any pancreatic metaplasia in any of our cohorts^26^. Previous reports have suggested that there is a distribution of gland type within the length of the BE segment^10^. While we observe a relationship between the individual gland types present at 1.0-2.0 cm and the overall size of the BE lesion, diversity of gland type is not dependent on lesion size, and cases where more than two phenotypes are present in the same biopsy are rare. This is confirmed when we investigate all biopsies taken at each particular endoscopy and we observe no change in gland phenotype diversity in the single biopsy compared with the diagnostic biopsies (**Supplemental Figure 2**).

Here, we report that gland phenotype can evolve and demonstrate that distinct gland types, each with a specific combination of differentiated epithelial cells, can share a common ancestry. In particular we show cardiac-type, specialized and mature intestinal glands all in direct proximity to each other have a common mtDNA mutation that can only be explained by the mutation arising in an ancestral gland and being passed to its daughter glands as it divides by gland fission^27^. The odds of two glands with distinct epithelial phenotypes possessing the same homoplasmic or highly heteroplasmic mtDNA mutation independently is vanishingly small^19^. It is therefore highly likely that gland fission is the mechanism of clonal expansion within the metaplastic esophagus as it is in the normal, unperturbed GI tract^19^. Importantly, we observed a shared somatic mtDNA mutation between oxynto-cardiac glands and specialized glands, demonstrating that intestinal and mature gastric lineages of differentiation can share ancestry. Although glandular differentiation within the BE segment is traditionally depicted as one of increasing intestinalization *per se*, these data demonstrate that metaplastic glands can also differentiate along a gastric line of differentiation (**Figure 2**).

Our clonal analysis data suggest that the cardiac-type gland is the fulcrum on which all other gland types observed in BE are based. Cardiac glands are characterized as simple glands, containing only foveolar cells at their surface and mucous secreting cells at their base. It has been suggested that the presence of CLO is associated with shorter lengths of BE and a lower cancer risk^28^. However, we have previously shown that CLO can evolve to cancer and can contain oncogenic driver mutations in genes such as TP53^7^. We therefore propose that evolution of cardiac-type glands is the initial basis of progression within BE.

The presence of multiple gland phenotypes across the BE segment may yield important information on the risk of progression to dysplasia or cancer. In terms of physical size, BE changes very little, if at all, over-time^29^. This belies the rate of clonal evolution within the BE segment itself, in particular those cases that are at risk of developing cancer^15^. Somatic genetic alterations and their diversity in particular are increased in BE prior to onset of cancer^14-16^. Controversy surrounds the evolution of genetic diversity with some studies demonstrating that this occurs only 2-4 years prior to the onset of cancer^16^ whereas others indicate that BE from patients who progress always have increased genetic diversity^14^. This applies to multiple cancer types^30^. Here, phenotypic diversity is shown to be increased in the non-dysplastic areas of Barrett’s surrounding dysplasia. We considered that analyzing biopsies may underestimate Shannon indices. Bootstrapping our sampling data however revealed that a stable Shannon index was achieved with the number of samples analyzed. While our data support the hypothesis that increased diversity of metaplastic phenotype adjacent to dysplasia reflects overall lesion diversity (as supported by our extensive data sampling of non-dysplastic biopsies in **Figure 1**), we were not able to collect sufficient specimen material to represent the entire length of the Barrett lesion in those patients that had undergone EMR. While our data cannot exclude the possibility that the presence of a dysplastic lesion secondarily drove the increased diversity in its neighboring mucosa, we consider this less likely. Indeed, data from the neo-Barrett patients suggest that no phenotypic diversity is present in recently developed Barrett and this is therefore acquired rather than inherent. Furthermore, in cases that progress to dysplasia, maximal genetic diversity has been shown to occur towards the gastroesophageal junction^31,32^. Our samples taken in this same region also demonstrate phenotypic diversity regardless of segment length. Phenotypic diversity in BE and other conditions^33^ in the progression to cancer is understudied and when we consider that tissue diagnoses rely on phenotypic analysis, it is surprising that diversity is not investigated more often.

Overall, this suggests that BE gland phenotypes are diverse, the presence of individual phenotypes are not related to the size of the lesion, and can be observed simultaneously in the same location within the esophagus. Our data shows definitively that BE phenotypes represent an evolutionary process and suggests that diversity of gland phenotype may play a role in progression.

## Data Availability

The datasets generated during and/or analysed during the current study are available from the corresponding author on reasonable request.

## Acknowledgements

We would like to thank the histology core service at the Barts Cancer Institute, Queen Mary University of London for providing all sectioning of tissues.

## Supplemental figure and table legends

**Supplemental Figure 1.** *Representative IHC for lineage identifying antibodies*. (A) Set −1, H+K+ATPase (DAB-brown) and pepsinogen (AP-blue). (B) Set −2. MUC5AC (DAB-brown) and MUC2 (AP-blue). (C) Set −3, Defensin 6 (AP-blue). Most biopsies were positive for MUC5AC and MUC2.

**Supplemental Figure 2.** *Individual patient distribution of gland phenotype based on available diagnostic tissue blocks*. All biopsies from a total of 25 patients, from the cohort described in Figure 1B (top), taken at the same endoscopy during which the single biopsy was taken, were phenotyped. H&E sections were reviewed by an expert pathologist (MJ) and two experienced researchers (EC and SMcD). Data is presented as percentage of each phenotype within each biopsy at the specific depth in the esophagus it was taken.

**Supplemental Figure 3.** *Diversity of BE gland phenotype is not based on patient age*. Each biopsy taken from figure 1B (top) was redistributed according to patient age at the time of endoscopy. There is no correlation between age of patient and phenotypic diversity.

**Supplemental Figure 4.** *Phenotypic richness in BE is not related to (A)patient age, (B)lesion size and (C) the number of biopsies taken at endoscopy*. The Spearman correlation coefficient was calculated between the number of biopsies sampled and the richness score for each of 1000 bootstrapping replicates for every patient (n=22). This shows that with more biopsies taken, the greater diversity is observed. Three patients were removed from this analysis due to unchanging richness across all biopsies.

**Supplemental Figure 5.** *Diversity on follow up is highly variable*. 10 patients with non-dysplastic BE show variable gland richness (no. phenotypes) over time. 5 patients showed no change in diversity, 3 showed an increase, 1 showed a decrease and 1 patient was variable. Red dot represents a change in phenotype with no change in richness.

**Supplemental Figure 6.** *Multiple gland phenotypes can show a common ancestor*. (**A**)MUC2 IHC revealing glands marked (i) MUC2-, ii) MUC2+ and iii) MUC2+. (**B**) A serial section stained for MUC5AC revealing glands (i) MUC5AC+, (ii) MUC5AC+ and (iii) MUC5AC-. Together gland (i) is cardiac type, (ii) is specialized and (iii) is mature intestinal. Each share a common somatic m.7148 T>C mutation in the MT-CO1 region of the mt genome (**C**) that was not present in the stroma (**D**).

**Supplemental Figure 7.** *The number of glands sampled in the BE and BE adjacent to dysplasia cohorts was sufficient to provide a reliable Shannon index result*. Each graph represents an individual patient. The blue shaded area represents the 95% confidence interval. Shannon index is derived from a bootstrapping subset (n=5000) from either the BE or the BE adjacent to dysplasia cohort. In the vast majority of cases a stable Shannon index was achieved in the simulation below or at the number of glands used in the experimental data set. Neo-Be was not included due to no diversity being observed.

**Supplemental Figure 8.** *Gland phenotype in Neo-BE cases*. Thirty-one biopsies from 19 Neo-B patient biopsies were phenotyped. The majority of patient biopsies revealed a pure cardiac phenotype.

**Supplement Figure 9.** A per-patient analysis of observed patient specimen gland phenotypes of non-dysplastic BE (Top left), adjacent to dysplasia BE (Top right) and neo-BE (Bottom) revealing phenotypic diversity within each group. Each column represents a single biopsy taken a known site within the esophagus.

**Supplemental Table 1.** Lineage-specific antibodies used to identify gland phenotype and their IHC conditions.

## Notes

### Competing Interest Statement

The authors have declared no competing interest.

### Author Declarations

London research ethics committee approval (11/LO/1613 and 15/LO/2127). County Durham and Tees Valley 2 research ethics committee approval (08/H0908/25)

## Bibliography

1. Pophali P, Halland M. Barrett’s oesophagus: diagnosis and management. BMJ 2016;353:i2373.

2. Shaheen NJ, Falk GW, Iyer PG, et al. ACG Clinical Guideline: Diagnosis and Management of Barrett’s Esophagus. Am J Gastroenterol 2016;111:30–50– quiz 51.

3. Fitzgerald RC, di Pietro M, Ragunath K, et al. British Society of Gastroenterology guidelines on the diagnosis and management of Barrett’s oesophagus. Gut 2014;63:7–42.

4. Bhat S, Coleman HG, Yousef F, et al. Risk of malignant progression in Barrett’s esophagus patients: results from a large population-based study. J Natl Cancer Inst 2011;103:1049–1057.

5. Kelty CJ, Gough MD, Van Wyk Q, et al. Barrett’s oesophagus: Intestinal metaplasia is not essential for cancer risk. Scand J Gastroenterol 2009;42:1271–1274.

6. Srivastava A, Golden KL, Sanchez CA, et al. High Goblet Cell Count Is Inversely Associated with Ploidy Abnormalities and Risk of Adenocarcinoma in Barrett’s Esophagus. PLoS ONE 2015;10:e0133403.

7. Lavery DL, Martinez P, Gay LJ, et al. Evolution of oesophageal adenocarcinoma from metaplastic columnar epithelium without goblet cells in Barrett’s oesophagus. Gut 2016;65:907–913.

8. McDonald SAC, Lavery D, Wright NA, et al. Barrett oesophagus: lessons on its origins from the lesion itself. Nature Publishing Group 2015;12:50–60.

9. Rhee H, Wang DH. Cellular Origins of Barrett’s Esophagus: the Search Continues. 2018:1–5.

10. Going JJ, Fletcher-Monaghan AJ, Neilson L, et al. Zoning of mucosal phenotype, dysplasia, and telomerase activity measured by telomerase repeat assay protocol in Barrett’s esophagus. Neoplasia 2004;6:85–92.

11. Paull A, Trier JS, Dalton MD, et al. The histologic spectrum of Barrett’s esophagus. N Engl J Med 1976;295:476–480.

12. Chandrasoma PT, Der R, Dalton P, et al. Distribution and significance of epithelial types in columnar-lined esophagus. Am J Surg Pathol 2001;25:1188–1193.

13. Lavery DL, Nicholson AM, Poulsom R, et al. The stem cell organisation, and the proliferative and gene expression profile of Barrett’s epithelium, replicates pyloric-type gastric glands. Gut 2014;63:1854–1863.

14. Martinez P, Timmer MR, Lau CT, et al. Dynamic clonal equilibrium and predetermined cancer risk in Barrett’s oesophagus. Nat Commun 2016;7:1–10.

15. Maley CC, Galipeau PC, Finley JC, et al. Genetic clonal diversity predicts progression to esophageal adenocarcinoma. Nat Genet 2006;38:468–473.

16. Li X, Galipeau PC, Paulson TG, et al. Temporal and spatial evolution of somatic chromosomal alterations: a case-cohort study of Barrett’s esophagus. Cancer Prevention Research 2014;7:114–127.

17. Reid BJ, Paulson TG, Li X. Accepted Manuscript. Gastroenterology 2015:1–28.

18. McDonald SAC, Graham TA, Lavery DL, et al. The Barrett’s Gland in Phenotype Space. JCMGH 2015;1:41–54.

19. Greaves LC, Preston SL, Tadrous PJ, et al. Mitochondrial DNA mutations are established in human colonic stem cells, and mutated clones expand by crypt fission. Proc Natl Acad Sci USA 2006;103:714–719.

20. Gutierrez-Gonzalez L, Deheragoda M, Elia G, et al. Analysis of the clonal architecture of the human small intestinal epithelium establishes a common stem cell for all lineages and reveals a mechanism for the fixation and spread of mutations. J. Pathol. 2009;217:489–496.

21. McDonald SAC, Greaves LC, Gutierrez-Gonzalez L, et al. Mechanisms of field cancerization in the human stomach: the expansion and spread of mutated gastric stem cells. Gastroenterology 2008;134:500–510.

22. Harrison R, Perry I, Haddadin W, et al. Detection of intestinal metaplasia in Barrett’s esophagus: an observational comparator study suggests the need for a minimum of eight biopsies. Am J Gastroenterol 2007;102:1154–1161.

23. Dunn LJ, Shenfine J, Griffin SM. Columnar metaplasia in the esophageal remnant after esophagectomy: a systematic review. Dis Esophagus 2015;28:32–41.

24. Shibata D. Inferring human stem cell behaviour from epigenetic drift. J. Pathol. 2009;217:199–205.

25. Baker A-M, Cereser B, Melton S, et al. Quantification of Crypt and Stem Cell Evolution in the Normal and Neoplastic Human Colon. Cell Reports 2014.

26. Krishnamurthy S, Dayal Y. Pancreatic metaplasia in Barrett’s esophagus. An immunohistochemical study. Am J Surg Pathol 1995;19:1172–1180.

27. Nicholson AM, Graham TA, Simpson A, et al. Barrett’s metaplasia glands are clonal, contain multiple stem cells and share a common squamous progenitor. Gut 2012;61:1380–1389.

28. Desai TK, Krishnan K, Samala N, et al. The incidence of oesophageal adenocarcinoma in non-dysplastic Barrett’s oesophagus: a meta-analysis. Gut 2012;61:970–976.

29. Cameron AJ, Lomboy CT. Barrett’s esophagus: age, prevalence, and extent of columnar epithelium. Gastroenterol. 1992;103:1241–1245.

30. Andor N, Graham TA, Jansen M, et al. Pan-cancer analysis of the extent and consequences of intratumor heterogeneity. Nature Medicine 2016;22:105–113.

31. Martinez P, Mallo D, Paulson TG, et al. Evolution of Barrett’s esophagus through space and time at single-crypt and whole-biopsy levels. Nat Commun 2018;9:794.

32. Timmer MR, Martinez P, Lau CT, et al. Derivation of genetic biomarkers for cancer risk stratification in Barrett’s oesophagus: a prospective cohort study. Gut 2016;65:1602–1610.

33. Almendro V, Kim HJ, Cheng Y-K, et al. Genetic and phenotypic diversity in breast tumor metastases. Cancer Res. 2014;74:1338–1348.

