## Supplemental methods for "Clonal transitions and phenotypic evolution in Barrett esophagus"

The Spearman correlation between richness of phenotype data and the number of biopsies was measured using the Hmisc package (v.4,3,0) in R, implementing a bootstrapping strategy. Each patient (index i) has n_i biopsies. A richness score was calculated by the presence of unique phenotypes over the set of biopsies, i.e. R(n_i). The richness R, was calculated (R(n_j)) for patient i, and a random subset of biopsies j, ranging from j=1,2,…,n_i. Each random subset obtained is a single bootstrap replicate and was repeated 1000 times. R(n_j) was calculated for each patient on their subsetted biopsies. The Spearman rank correlation coefficient of R(n_i) was calculated between the number of biopsies sampled and the richness score R(n_j) for each of the 1000 replicates for each patient. The final coefficient values across 22 patients (excluding patients with unchanging richness across all biopsies) were ranked from low to high prior to visualization using the ggplot2 package (v.3.2.1) in R.

To ensure sufficient sampling was achieved for diversity per biopsy between non-dysplastic BE and BE adjacent to dysplasia, biopsies from each disease state were independently sub-sampled 1000 times with a random number of glands in each subset (each subset being a bootstrap). Each individual patient data was plotted separately.
