## supplemental table 1 for "Clonal transitions and phenotypic evolution in Barrett esophagus"

**Characteristics of the primary antibodies used for IHC**

| Antibody | Source | Host/Isotype | Dilution FFPE | Dilution FF | Secondary | Tertiary |
|---|---|---|---|---|---|---|
| Pepsinogen | Abcam | Mouse/IgG1 | 1:700 | N/A | Goat anti mouse | AP/Blue |
| H+K+ATPase | DAKO | Mouse/IgG1 | 1:1000 | 1:4000 | Goat anti mouse | HRP/DAB |
| MUC5AC | Abcam | Mouse/IgG1 | 1:500 | 1:50/ 1:100 | Goat anti mouse | HRP/DAB |
| MUC2 | ThermoFisher Scientific | Mouse/IgG1 | 1:500 | 1:200 | Goat anti mouse | AP/Blue |
| Defensin6$\alpha$ | Sigma | Rabbit/ Polyclonal | 1:5000 | N/A | Swine anti rabbit | AP/Blue |
| Defensin5$\alpha$ | Abcam | Mouse/IgG1 | N/A | 1:100 | Rabbit anti mouse | HRP/DAB |

Supplemental table 1. Antibody sources, dilutions and processing used within this manuscript. FFPE= formalin fixed paraffin embedded, FF= fresh frozen and N/A = not applicable; antibody not used for this purpose. AP= alkaline phosphatase and HRP= horseradish peroxidase.
